# Recruitment for dementia clinical trials in care homes: an evaluation of strategies employed in the Sativex® for the treatment of Agitation in Dementia (‘STAND’) Trial

**DOI:** 10.1101/2025.06.25.25330292

**Authors:** Christopher P Albertyn, Byron Creese, Ta-Wei Guu, Simrat Kaur, Pooja Kandangwa, Miguel Da Silva, Dag Aarsland

## Abstract

Clinical trials in dementia face significant recruitment challenges, with only 1% of eligible participants typically engaged in research. The ‘Sativex for the Treatment of AgitatioN in Dementia’ (STAND) trial implemented innovative recruitment strategies to address these barriers. This study analyses the effectiveness of systematic recruitment approaches combining public outreach, targeted engagement, and electronic consent procedures. A mixed-methods approach incorporated patient and public involvement (PPI), stakeholder mapping, and iterative recruitment channel optimisation. Over 10 months, 98 participant enquiries were received, resulting in 53 participants consented and 29 enrolled (55% screen success rate). Electronic consent emerged as the preferred method (49% of consents), reducing time from first contact to signed informed consent from approximately 34 days (in-person/postal) to just 5 days. Pre-existing research networks provided 83% of participants, demonstrating their value. Despite falling short of the target 60 participants, primarily due to COVID-19 restrictions and drug supply challenges, recruitment exceeded targets in later months following implementation of PPI-informed strategies and protocol amendments. The study demonstrates that systematic recruitment approaches incorporating eConsent can effectively accelerate enrolment in dementia trials, while highlighting the importance of flexibility in protocol design and the value of embedded clinical research infrastructure within healthcare systems.

**Key points:**

- Systematic, stakeholder-driven recruitment strategies improved care home dementia trial enrolment rates
- Electronic consent reduced time to consent from 34 days (in-person/postal) to just 5 days
- Pre-existing research networks provided 83% of participants, demonstrating their critical value
- Protocol flexibility and PPI enabled rapid adaptation to COVID-19 and operational challenges
- Combining eConsent, embedded infrastructure, and adaptive management overcame recruitment barriers

## Introduction

Clinical trials are the cornerstone of developing effective interventions for Alzheimer’s disease (AD) and other dementias, yet they consistently face significant recruitment challenges that exceed those encountered in most other therapeutic areas (1). These studies typically require longer recruitment periods, incur higher costs, and achieve lower positive screen rates compared to trials in other disease domains (2). Despite substantial resources invested in dementia research, the field continues to struggle with "Lasagna’s Law" – the phenomenon where patient availability sharply decreases when a clinical trial begins (3).

The recruitment challenge is particularly striking: only 1% of potentially eligible participants are typically engaged by the clinical trial ecosystem (4), and merely 2% of individuals diagnosed with dementia in the UK join dementia research registries (5). Multiple factors contribute to these difficulties, including limited physician time and diagnostic tools, dementia stigma, study partner requirements, and complex consent procedures for advanced dementia patients involving legally authorised representatives (6,7).

Various strategies have been proposed to address these challenges, including increasing public and clinical staff engagement, enhancing disease-specific education, creating integrated "research-ready" clinical environments, and optimising recruitment team capacity (4). However, the systematic implementation and evaluation of these approaches within the context of specific trials remains limited, creating a significant knowledge gap regarding their effectiveness.

The Sativex for the Treatment of AgitatioN in Dementia (’STAND’) trial provides an opportunity to address this gap. This double-blind, placebo-controlled feasibility trial investigated a cannabinoid-based oral spray for treating agitation in UK nursing home residents with Alzheimer’s Disease (8,9). The trial incorporated novel recruitment strategies specifically designed to overcome known barriers in the dementia trial space.

This paper describes and evaluates these innovative approaches, including systematic stakeholder mapping, dynamic engagement strategies, and electronic consent procedures, to provide evidence-based insights for future dementia clinical trials.

## Methods

### Study Design and Setting

The STAND trial was a double-blind, placebo-controlled feasibility study investigating Sativex®, a cannabinoid-based oral spray, for treating agitation in nursing home residents with Alzheimer’s disease. The study aimed to recruit 60 participants from care homes within Greater London over 12 months, targeting 5 participants per month. The trial was approved by relevant ethics committees (ISRCTN registry 7163562). A full list of the eligibility criteria is included in the supplementary materials, the published protocol and the main outcomes papers (8,9).

### Novel Recruitment Strategy Development

Building on previous research recommendations (1), we embedded a systematic, dynamic recruitment strategy informed by patient and public involvement (PPI), incorporating four core elements:

#### 1. Dynamic Multistakeholder Mapping and Engagement

We used Mendelow’s Power-Interest Matrix (10) to systematically map and prioritise diverse actors influencing recruitment outcomes. Empirical evidence demonstrates stakeholder-driven strategies can increase dementia trial enrolment.. Empirical evidence demonstrates stakeholder-driven strategies can increase dementia trial enrolment, particularly when involving persons living with dementia (PLWD) and caregivers in protocol design (11,12). Stakeholders - who have influence on recruitment outcomes (see supplementary materials for full list) - were categorised into four quadrants:

- ‘Key Players’ (high influence, high interest): These stakeholders are crucial to the project’s success. They require regular, detailed communication and active engagement (e.g. the Care Home Research Network (CHRN) and dedicated dementia PPI groups).
- ‘Manage Closely’ (high influence, low interest): These stakeholders have significant influence but are less concerned with day-to-day activities. They need to be kept satisfied with high-level updates (e.g. community healthcare teams, primary care physicians, and Care Quality Commission-listed care homes).
- ‘Keep Informed’ (low influence, high interest): These stakeholders are interested in the project but lack the power to influence it. Regular updates and information sharing are essential (e.g. care home media outlets and NIHR Clinical Research Networks).
- ‘Monitor’ (low influence, low interest): These stakeholders require minimal effort. They should be monitored to ensure they remain supportive or neutral (e.g. academic colleagues and patient recruitment registries).

For each channel, we specified the function of engagement (direct recruitment, PPI advice, education, or dissemination), appropriate contact methods, frequency of contact, and assigned study personnel to ‘take point’, ensuring that resources were shared and managed effectively. This "living document" was updated monthly based on feedback and recruitment outcomes whereby stakeholders could move between categories depending on their current level of power and/or interest and the recruitment environment (e.g. adapting to covid restrictions). A visual outline of the matrix is provided in Figure 1. The matrix’s strength lies in its dynamic adaptability, enabling us to pivot our recruitment strategy, resources and efforts using a data-driven and locally PPI informed approach. This iterative approach counters static strategies that fail to address the evolving landscape of recruitment barriers and facilitators. Instructions for operationalising this matrix are outlined in the supplementary materials.

**Figure 1:**
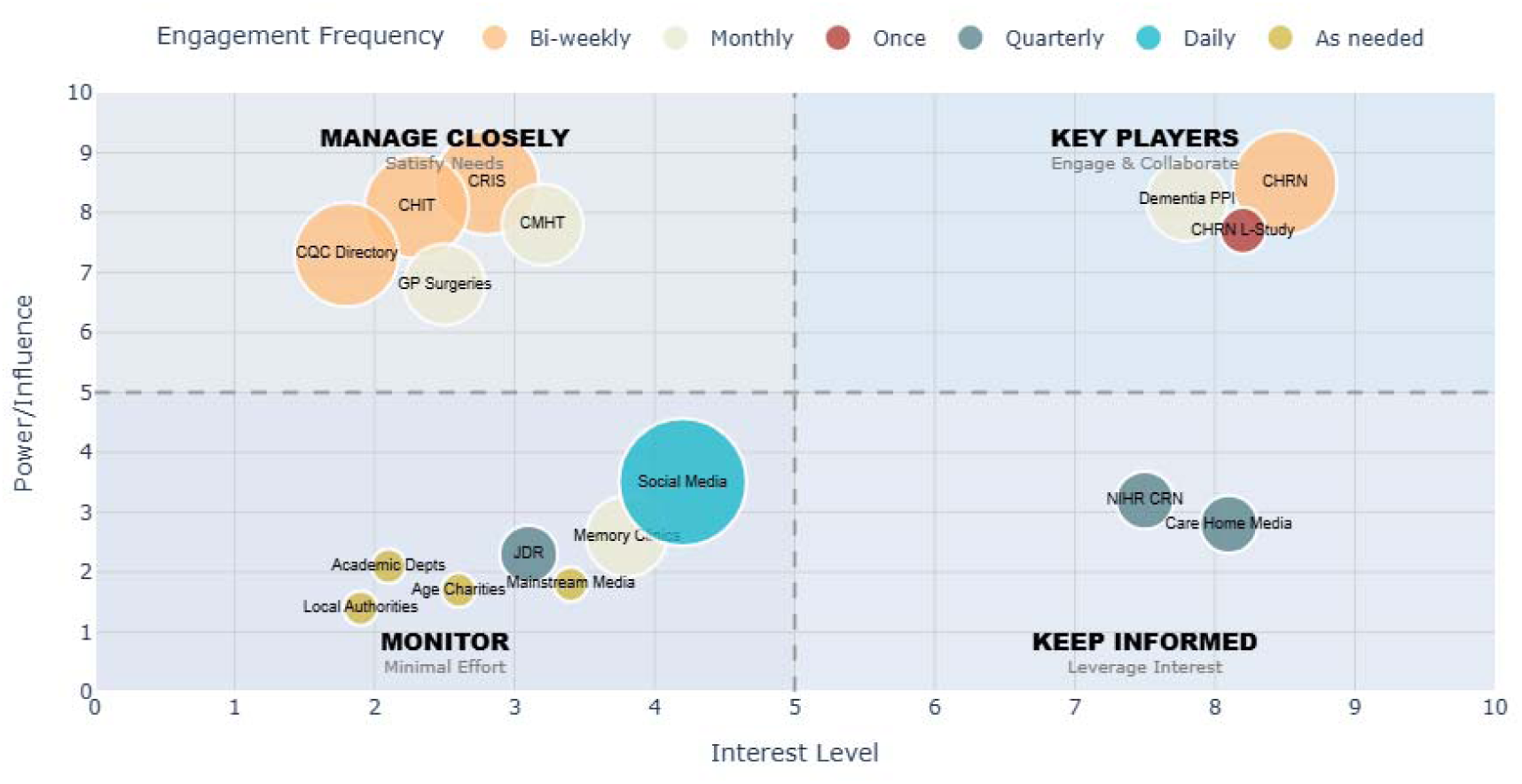
STAND Trial multistakeholder recruitment channel mapping matrix. Recruitment channels are positioned according to their influence on recruitment outcomes (power) and their level of engagement (interest), with marker size reflecting frequency of contact. Quadrants indicate recommended engagement strategy: ‘Key Players’ (high power, high interest) require intensive collaboration; ‘Manage Closely’ (high power, low interest) need regular engagement; ‘Keep Informed’ (low power, high interest) benefit from information sharing; and ‘Monitor’ (low power, low interest) require minimal oversight. This adaptive mapping guided targeted, efficient resource allocation and dynamic engagement throughout the trial

#### 2. Optimised Consenting Protocol

Recognising consent complexities for advanced dementia patients (13,14), we developed a systematic approach offering multiple pathways:

- Direct participant consent (when capacity present)
- Personal Legal Representative (PerLR) consent (family/friends)
- Professional Legal Representative (ProLR) consent (care home/clinical staff) To maximise accessibility and efficiency, we offered three consent modalities:
- Traditional in-person consent.
- Postal consent with prepaid return envelopes.
- Electronic consent (eConsent) via Adobe Sign.

Three consent modalities were offered: traditional in-person, postal with prepaid envelopes, and electronic consent (eConsent) via Adobe Sign, implemented following HRA and MHRA guidance (15). For a flow diagram of the optimised consenting protocol, see supplementary materials.

#### 3. PPI-Informed Multimedia Approach

Monthly PPI engagement sessions with both patient groups and care home partners informed the development of recruitment materials and strategies. Based on PPI feedback, we created:

- Simplified information sheets tailored to cognitive capacity levels
- Animated explanatory videos for those with reading difficulties (16)
- Targeted messaging avoiding stigmatizing language
- Educational workshops for care home staff

See the following online repository for some of the resources used in the trial (17).

#### 4. Adaptive Protocol Management

Continuous feedback mechanisms identified barriers and implemented mitigating strategies through recruitment trend analysis, stakeholder feedback sessions, and protocol amendment planning.

### Data Collection and Analysis

We collected quantitative data on recruitment sources, consent methods, time to consent, and monthly recruitment rates. Qualitative insights were gathered through PPI engagement sessions and stakeholder feedback.

## Results

### Recruitment channels identified from multistakeholder mapping and engagement activities

Through iterative multistakeholder mapping, 17 recruitment channels were identified (see supplementary materials). Each channel was categorised within the priority matrix and assigned specific trial staff members. This "living document" adapted throughout the trial based on changing circumstances. Key channels included:

- Care Home Research Network (CHRN): Connects >200 care homes in London/southeast England
- Clinical Records Interactive Service (CRIS): Secure NHS database enabling targeted participant identification
- NIHR Clinical Research Networks: National networks supporting research delivery
- Social Media/Care Home Media: Broad outreach and professional awareness Recruitment Overview

The recruitment window was reduced to 10 months (September 2021-June 2022) due to COVID-19 delays and drug supply constraints. **98 participant enquiries** meeting primary inclusion criteria were received, with **29 care homes** expressing interest.

#### Key outcomes

- **53 participants** consented and screened
- **29 participants** enrolled (55% screen success rate)
- **18 care homes** signed participation agreements
- Primary reasons screen failures: COVID-related exclusions (n=9), non-AD dementia diagnosis (n=8) (see main outcomes paper for full review of screen failures (9).

Recruitment rates remained below target between September 2021-March 2022 but exceeded targets in May-June 2022 (see Figure 2).

**Figure 2:**
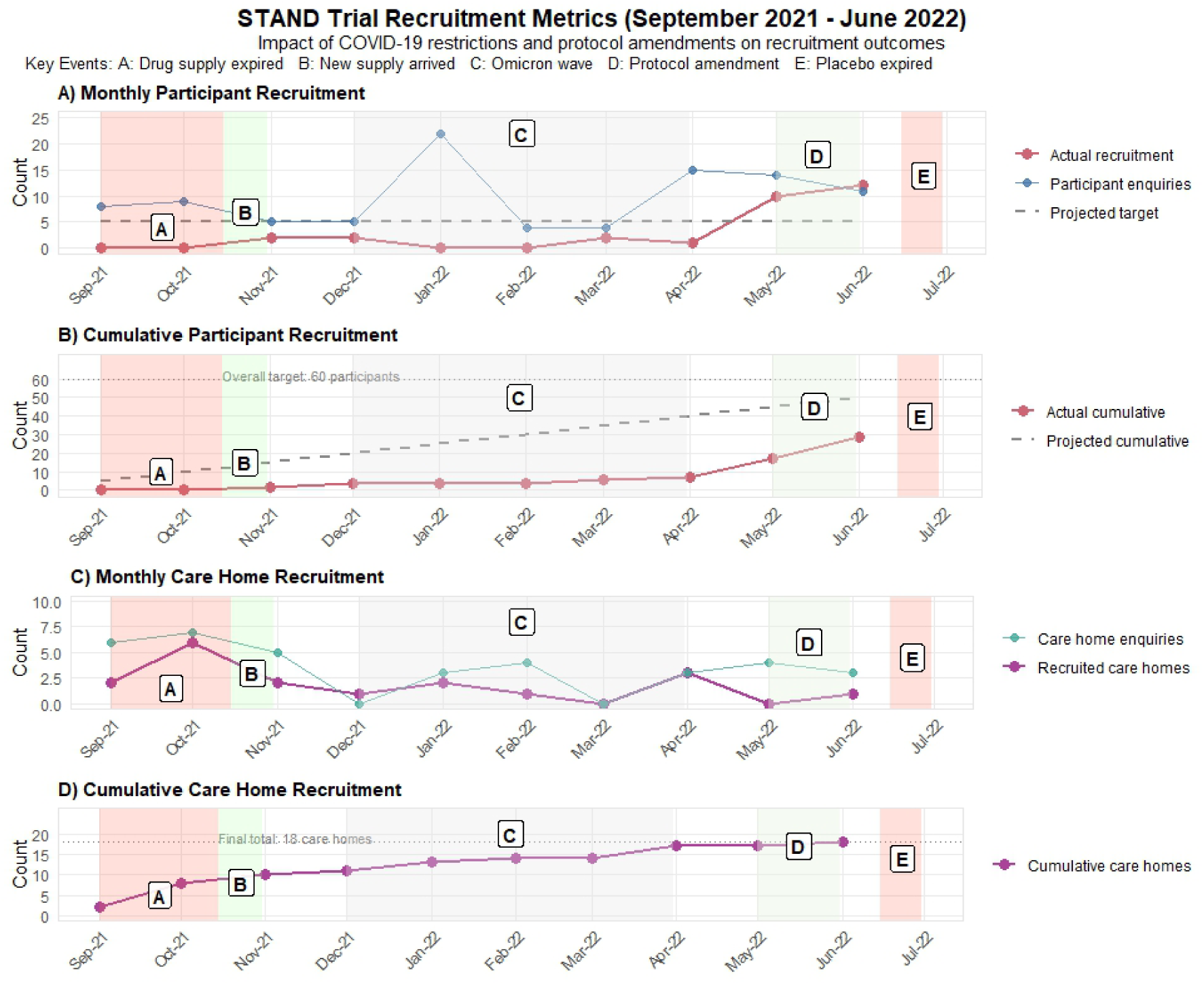
STAND Trial Recruitment Metrics. Recruitment dynamics in the STAND trial (September 2021-June 2022). This four-panel visualisation tracks participant and care home recruitment alongside critical operational events. Panels A and C show monthly recruitment rates for participants and care homes respectively, while Panels B and D display their cumulative totals against targets. Colour-coded markers highlight key events affecting recruitment: (A) initial drug expiry delaying start, (B) new drug supply arrival, (C) Omicron variant covid-19 wave restricting care home access, (D) protocol amendment reducing in-person procedures, and (E) placebo expiry necessitating early closure. The figure demonstrates how external challenges and strategic adaptations influenced recruitment patterns, with rates exceeding monthly targets after protocol amendments despite the shortened recruitment window.

### Consent Methods and Sources

Analysis of consent methods revealed that electronic consent (eConsent) was the most frequently used approach, accounting for 49% of all consents obtained (Table 1). Traditional in-person and postal consent methods were used for 30% and 21% of participants, respectively. Notably, eConsent significantly reduced the time between first contact and obtaining signed informed consent, averaging just 5.19 days (SD=4.00) compared to 33.69 days (SD=16.36) for in-person and 34.73 days (SD=15.17) for postal methods (Table 2).

**Table 1:** Describing source and method of signed consent forms.

| Consent form Signee | Method of signing |  |  | Total (%) |
| --- | --- | --- | --- | --- |
|  | In-person | Postal | eConsent |  |
| Participant | 0 | 0 | 0 | 0 (0) |
| Personal Legal Representative | 4 | 10 | 19 | 33 (62) |
| Professional Legal Representative | 12 | 1 | 7 | 20 (38) |
| Total (%) | 16 (30) | 11 (21) | 26 (49) | 53 |

\*Number represents number of participants (%) approached for consent to start screening

**Table 2:** Describing average number of days between 1st contact and signed ICF.

| Method of signing | Average no. of days between 1st contact and signed ICF (SD) |
| --- | --- |
| In-person | 33.69 (16.36) |
| Postal | 34.73 (15.17) |
| eConsent | 5.19 (4.00) |

\*Number represents the mean number of days (standard deviation) between contacting a Signee to a fully executed signed consent form.

Regarding consent sources, approximately two-thirds (62%) of consent forms were signed by Personal Legal Representatives (family members or friends), while the remaining third (38%) were provided by Professional Legal Representatives (healthcare professionals) (see Table 2). No participants were deemed to have capacity to consent directly for themselves, which is consistent with the advanced dementia population targeted by the study.

### Recruitment Channels

The majority of participants (83%) were identified through pre-existing "research-ready" clinical databases, networks, and established care home partnerships. The Care Home Research Network (CHRN) provided 45% of participants, while the NIHR Clinical Research Network and the Clinical Record Interactive Service (CRIS) contributed 20% and 18%, respectively. The remaining 17% came from other sources, including direct care home contact, social media outreach, and community mental health teams (see Figure 3).

**Figure 3:**
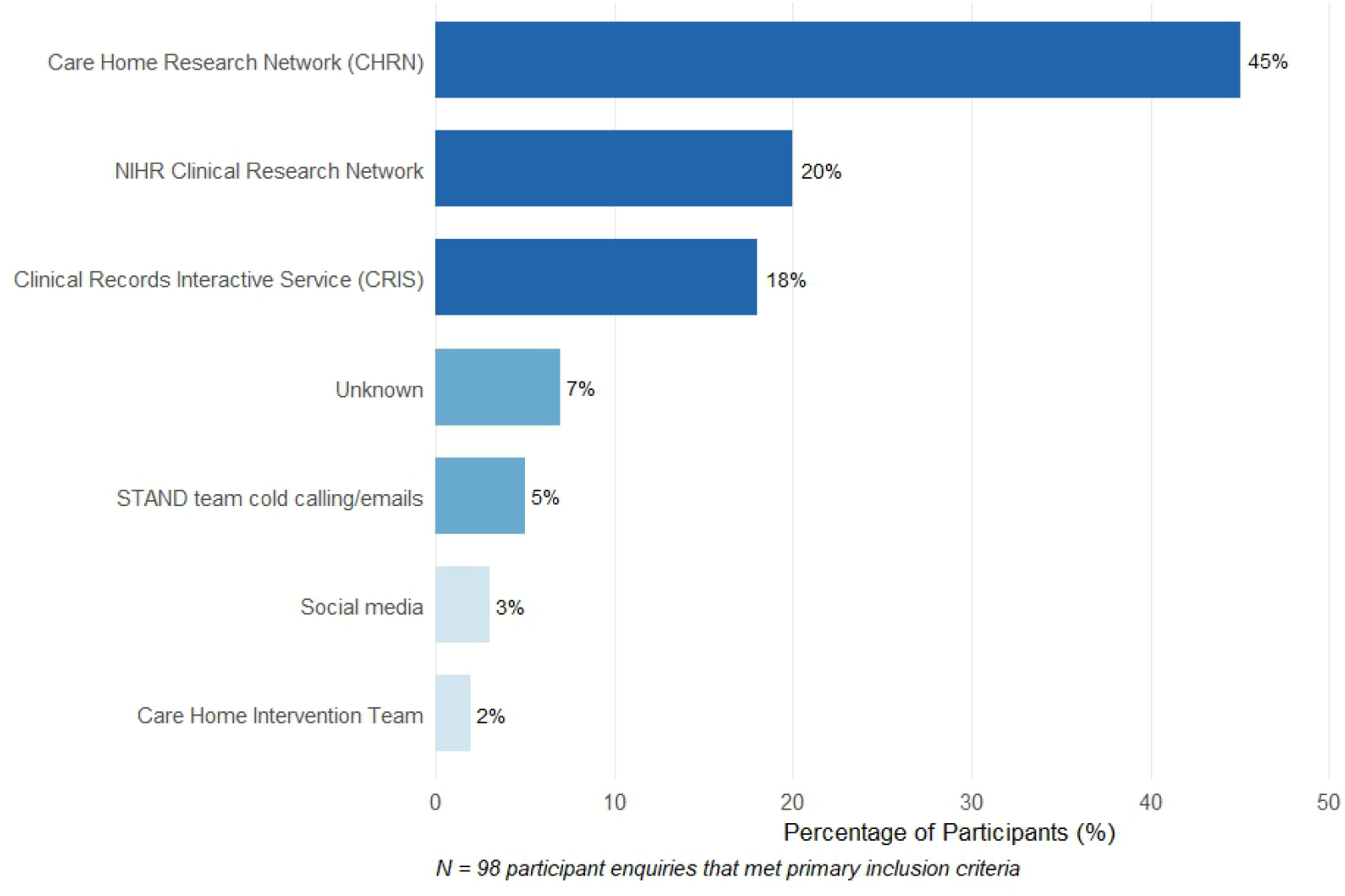
Recruitment Channel Effectiveness. The horizontal bar chart illustrates the distribution of participant identification sources (N=98) in the STAND trial. Pre-existing research infrastructure accounted for 83% of all referrals, with the Care Home Research Network providing nearly half (45%) of all participants, followed by the NIHR Clinical Research Network (20%) and Clinical Records Interactive Service (CRIS) (18%). Direct outreach methods such as cold calling (5%), social media (3%), and care home intervention teams (2%) produced significantly fewer enquiries, highlighting the critical value of embedded research networks for dementia trial recruitment in care home settings.

### Impact of External Events and Interventions

Several key events significantly influenced recruitment rates throughout the study:

- **Drug Supply Challenges** (September-October 2021): Initial recruitment was delayed due to drug supply expiration, with randomisation commencing only after new supply arrived in mid-October 2021.
- **COVID-19 Omicron Wave** (December 2021-March 2022): The emergence of the Omicron covid variant resulted in restricted access to care homes, significantly impacting recruitment during this period.
- **Protocol Amendment** (February-May 2022): A substantial amendment to reduce reliance on in-person invasive procedures was submitted in response to care home concerns about COVID-19 transmission. While this amendment took three months to receive approval, its implementation in May 2022 corresponded with a significant increase in recruitment.
- **PPI and Education Activities**: Notable spikes in recruitment enquiries followed specific PPI and education activities, including a care home workshop providing up to date evidence on the management of behavioural and psychological symptoms of dementia in September 2021, a care provider podcast in February 2022, and clinical network engagement sessions (within the CHRN, CHIT, CMHT and other clinical groups).
- **Drug Supply Expiration** (June 2022): The trial’s recruitment window was prematurely closed due to placebo product expiration, despite continued interest from potential participants.

## Discussion

This study demonstrates both the challenges and potential solutions in recruiting participants for dementia clinical trials, particularly within care home settings. Despite falling short of the target 60 participants, our systematic approach yielded important insights about effective recruitment strategies in this complex environment.

### Effectiveness of Novel Recruitment Approaches

The systematic stakeholder mapping and dynamic engagement strategy proved valuable in identifying and accessing diverse recruitment channels. The approach allowed for targeted resource allocation and relationship building with key stakeholders, consistent with previous recommendations emphasising the importance of multistakeholder engagement in dementia research (4,18).

The most striking finding was electronic consent’s superior performance, which significantly reduced time to obtain informed consent compared to traditional methods. Reduction from approximately 34 days to just 5 days represents a substantial improvement in recruitment efficiency. This finding aligns with emerging evidence supporting eConsent in clinical trials (19,20), particularly for studies involving participants with cognitive impairment who rely on legally authorised representatives who may not be physically present at the research site.

The value of "research-ready" infrastructure embedded within healthcare systems was clearly demonstrated, with pre-existing networks providing 83% of participants. The CHRN demonstrated significant value by recruiting approximately half of participants, fundamentally challenging traditional recruitment paradigms by providing immediate mobilisation capabilities that eliminated delays typically associated with cold-calling approaches. The network’s pre-established framework and trust proved invaluable during the COVID-19 pandemic, enabling continued engagement when care homes faced unprecedented restrictions, affording us the ability and knowledge to pivot and adapt our study protocol to continue study recruitment safely.

The CHRN’s "research-ready" care home cohort allowed recruitment efforts to focus on study-specific eligibility rather than fundamental willingness to participate. This difference was salient when recruiting care homes that were not CHRN members via CQC or other sources. These ‘research-naïve’ care homes required more intensive engagement and training before agreeing to participate. This infrastructure model highlights the value of shifting from resource-intensive, project-based recruitment to a sustainable, platform/infrastructure-based approach that creates continuous learning opportunities and institutional memory, ultimately demonstrating how embedded research networks can accelerate recruitment timelines, reduce costs, and improve study efficiency across the broader research ecosystem.

The effectiveness of the Clinical Records Interactive Service (CRIS) in pre-screening potential participants based on electronic health records highlights the potential of leveraging digital health data to optimise recruitment. This reinforces the importance of sustainable research infrastructure within clinical settings, as advocated by several experts in the field (4,21,22).

### Barriers and Adaptations

The study faced significant external barriers, most notably the COVID-19 pandemic, which restricted access to care homes during the Omicron wave. This experience echoes challenges reported across the clinical trial landscape during the pandemic, with particular impact on vulnerable populations (23). Our response - a protocol amendment reducing reliance on in-person procedures - demonstrated the importance of flexibility in trial design and value for responsive adaptations based on stakeholder feedback.

Drug supply challenges represented another significant barrier, highlighting logistical complexities that can impact recruitment timelines. Better tracking of product shelf-life and earlier planning for supply renewals would have mitigated these issues.

Notably, recruitment improved substantially following the protocol amendment and coinciding with reduced COVID-19 restrictions, suggesting that with continued operation, the trial might have approached or met its original recruitment target. The pronounced increase in recruitment rates in May-June 2022 (exceeding the target of 5 participants per month) provides evidence that the systematic approach was gaining effectiveness over time.

### Implications for Future Research

Several key learnings from this study have implications for future dementia clinical trials:

1. **Electronic consent should be prioritised**, particularly for studies involving participants with cognitive impairment who rely on legally authorised representatives. The substantial time savings and apparent preference for this method among signatories suggest it could significantly improve recruitment efficiency. The National Institute on Aging has actively promoted electronic consent pilot programmes since 2017, specifically targeting Alzheimer’s disease research studies, bringing together clinical researchers and data scientists to develop processes that remove key recruitment barriers (24). In the UK, the Health Research Authority and MHRA published joint guidance in 2018 supporting electronic consent methods, confirming that eConsent can supplement or replace traditional paper-based approaches whilst improving understanding and reducing dropout rates (15). UK academic trials units have demonstrated successful implementation of eConsent across diverse populations, including older adults with complex conditions, with one study showing 31% uptake amongst consented participants and no age-related barriers to adoption (25).
2. **Investment in research-ready infrastructure within healthcare systems** appears to yield substantial benefits for recruitment as demonstrated by the success of the CHRN. Expanding access across healthcare trusts and breaking down cross-organisational barriers could further enhance recruitment capacity. Previous initiatives such as the UK’s DeNDRoN (Dementias and Neurodegenerative Diseases Research Network) and ENRICH (Enabling Research in Care Homes) network also demonstrated this approach’s effectiveness. The DeNDRoN network, increased NHS sites participating in dementia studies from 31 to 188 sites between 2007-2011, with patient recruitment rising from 911 to 6,700 participants (26). The ENRICH (Enabling Research in Care Homes) network established research-ready care home infrastructure across England, Wales, and Scotland, facilitating recruitment whilst providing ongoing support and training to care home staff (27). The NIHR Dementia Translational Research Collaboration (Dementia TRC) is a UK-based initiative that will bring together leading dementia research centres to accelerate the translation of discoveries into clinical practice, improving diagnosis and treatment through collaboration with industry, academics, and healthcare partners (28,29).
3. **Public engagement and education activities** demonstrably increase recruitment enquiries. Strategic timing of these activities throughout the recruitment period may help maintain consistent interest and enrolment. Join Dementia Research, the UK’s national dementia research registry, has registered over 91,000 people and facilitated 94,651 participants joining studies across 334 research sites, demonstrating the power of coordinated public engagement (30). Alzheimer’s Research UK’s Inspire Fund specifically targets underserved communities through £5,000 grants for public engagement projects (31), whilst initiatives like Dementias Platform UK’s "Your Beautiful Brain" workshops have successfully engaged Black African and Caribbean communities who are traditionally under-represented in dementia research (32).
4. **Protocol flexibility** is essential, particularly in vulnerable populations and unpredictable circumstances. Building adaptability into study designs (e.g. including adaptive trial designs, different recruitment pathway options, different methods of assessment data collection) from the outset may improve resilience to external challenges. Adaptive trial designs allow learning from accumulating data and application of insights within ongoing trials without undermining validity, enabling modifications to doses, sample size, and entry criteria based on pre-specified response patterns (33–35). The BAN2401 (lecanemab) Phase 2 trial successfully used Bayesian dose-adaptive methodology to identify optimal dosing whilst adapting randomisation ratios during interim analyses, though such adaptive approaches remain underutilised in UK dementia trials (36).
5. **Cross-organisational collaboration** should be strengthened to overcome institutional barriers. Fragmentation of healthcare systems and research networks creates artificial boundaries that impede efficient participant identification and recruitment. UK research has identified that fragmented dementia services result in poor communication between providers and long gaps between services, with 49% of people with dementia receiving insufficient post-diagnostic support due to systemic fragmentation (37). Alzheimer’s Research UK has highlighted how the UK’s fragmented healthcare system creates suboptimal links between clinical research and dementia diagnoses in the NHS, recommending the establishment of a national clinical trials network to provide critical mass of expertise and break down organisational silos (5).

### Limitations

This study has several limitations. The lack of a control condition and concurrent implementation of multiple recruitment strategies makes it difficult to isolate effects of single approaches. The extraordinary circumstances of COVID-19 may limit generalisability to non-pandemic contexts. The restricted geographic focus on Greater London may not reflect recruitment challenges in other regions, particularly rural areas with different healthcare infrastructure.

### Conclusion

Despite not achieving the original recruitment target, this study demonstrates that systematic recruitment approaches incorporating electronic consent can effectively accelerate enrolment in dementia clinical trials. The substantial increase in recruitment following protocol adaptations suggests that with continued operation, the approach could have achieved or exceeded targets.

Our findings provide empirical validation for previously advocated recruitment strategies, demonstrating that systematic stakeholder engagement, research-ready infrastructure, and enhanced clinical partnerships can substantially improve recruitment outcomes in real-world dementia trials. However, rigorous controlled evaluation of individual strategy components remains necessary to optimise their implementation across diverse clinical contexts.

Future research should focus on refining these methods, particularly expanding electronic consent options, strengthening research-ready infrastructure across healthcare systems, and developing more robust contingency planning for external disruptions. Implementation of these strategies could help address the persistent challenge of slow recruitment in dementia research, ultimately accelerating development of interventions for this growing public health priority.

## Data Availability

All data produced in the present study are available upon reasonable request to the authors.

## References

1. Dana Goldman P. Key Barriers for Clinical Trials for Alzheimer’s Disease. 2020 Aug 17 [cited 2023 Jan 22]; Available from: https://healthpolicy.usc.edu/research/key-barriers-for-clinical-trials-for-alzheimers-disease/

2. Cummings J, Ritter A, Zhong K. Clinical Trials for Disease-Modifying Therapies in Alzheimer’s Disease: A Primer, Lessons Learned, and a Blueprint for the Future1. J Alzheimers Dis. 64(Suppl 1):S3–22.

3. Bogin V. Lasagna’s law: A dish best served early. Contemporary Clinical Trials Communications. 2022 Apr 1;26:100900.

4. Langbaum JB, Zissimopoulos J, Au R, Bose N, Edgar CJ, Ehrenberg E, et al. Recommendations to address key recruitment challenges of Alzheimer’s disease clinical trials. Alzheimer’s & Dementia. 2023;19(2):696–707.

5. Alzheimer’s Research UK [Internet]. [cited 2023 Jan 22]. Translating Science into breakthroughs: the future of late-stage dementia clinical trials in the UK. Available from: https://www.alzheimersresearchuk.org/about-us/our-influence/policy-work/reports/translating-science-into-breakthroughs-the-future-of-late-stage-dementia-clinical-trials-in-the-uk/

6. Study partners perform essential tasks in dementia research and can experience burdens and benefits in this role - Betty S Black, Holly A Taylor, Peter V Rabins, Jason Karlawish, 2018 [Internet]. [cited 2025 May 28]. Available from: https://journals.sagepub.com/doi/10.1177/1471301216648796

7. Mozersky J, Solomon ED, Baldwin K, Wroblewski M, Parsons M, Goodman M, et al. Barriers to Using Legally Authorized Representatives in Clinical Research with Older Adults. J Alzheimers Dis Rep. 2023;7(1):135–49.

8. Albertyn CP, Guu TW, Chu P, Creese B, Young AH, Velayudhan L, et al. Sativex (Nabiximols) for the treatment of Agitation &amp; Aggression in Alzheimer’s Dementia in UK nursing homes (STAND): a randomised, double-blind, placebo-controlled feasibility trial [Internet]. medRxiv; 2024 [cited 2025 May 12]. p. 2024.12.18.24319032. Available from: https://www.medrxiv.org/content/10.1101/2024.12.18.24319032v1

9. Albertyn CP, Guu TW, Chu P, Creese B, Young A, Velayudhan L, et al. Sativex (nabiximols) for the treatment of Agitation & Aggression in Alzheimer’s dementia in UK nursing homes: a randomised, double-blind, placebo-controlled feasibility trial. Age and Ageing. 2025 Jun 1;54(6):afaf149.

10. Barrington R. What Is Mendelow’s Matrix And How Is It Useful? [Internet]. Oxford College of Marketing Blog. 2018 [cited 2025 Jun 2]. Available from: https://blog.oxfordcollegeofmarketing.com/2018/04/23/what-is-mendelows-matrix-and-how-is-it-useful/

11. Patel NK, Masoud SS, Meyer K, Davila AV, Rivette S, Glassner AA, et al. Engaging multi-stakeholder perspectives to identify dementia care research priorities. J Patient Rep Outcomes. 2021 Jun 22;5:46.

12. Hirt J, Beer T, Cavalli S, Cereghetti S, Pusterla ERG, Zeller A. Recruiting Persons With Dementia: A Systematic Review of Facilitators, Barriers, and Strategies. Am J Alzheimers Dis Other Demen. 2024 Aug 13;39:15333175241276443.

13. Evans CJ, Yorganci E, Lewis P, Koffman J, Stone K, Tunnard I, et al. Processes of consent in research for adults with impaired mental capacity nearing the end of life: systematic review and transparent expert consultation (MORECare_Capacity statement). BMC Med. 2020 Jul 22;18:221.

14. Diaz A, Birck C, Bradshaw A, Georges J, Lamirel D, Moradi-Bachiller S, et al. Informed consent in dementia research: how Public Involvement can contribute to addressing “old” and “new” challenges. Front Dement. 2025 Feb 4;4:1536762.

15. Health Research Authority [Internet]. [cited 2020 Apr 15]. HRA and MHRA publish joint statement on seeking and documenting consent using electronic methods (eConsent). Available from: /about-us/news-updates/hra-and-mhra-publish-joint-statement-seeking-and-documenting-consent-using-electronic-methods-econsent/

16. (1) Facebook [Internet]. [cited 2025 Jun 16]. Available from: https://www.facebook.com/standtrialkcl/videos/2811607975752669

17. Albertyn C. Study materials, SPIRIT checklist and full Statistical Analysis Plan for the Sativex® for the treatment of Agitation & Aggression in Alzheimer’s Dementia in UK nursing homes (’STAND’) trial. 2023 May 30 [cited 2024 May 30]; Available from: https://figshare.com/articles/journal_contribution/Study_materials_SPIRIT_checklist_and_full_Statistical_Analysis_Plan_for_the_Sativex_for_the_treatment_of_Agitation_Aggression_in_Alzheimer_s_Dementia_in_UK_nursing_homes_STAND_trial_/23260934/1

18. Nocivelli B, Shepherd V, Hood K, Wallace C, Wood F. Identifying barriers and facilitators to the inclusion of older adults living in UK care homes in research: a scoping review. BMC Geriatrics. 2023 Jul 20;23(1):446.

19. Jaton E, Stang J, Biros M, Staugaitis A, Scherber J, Merkle F, et al. The Use of Electronic Consent for COVID-19 Clinical Trials: Lessons for Emergency Care Research During a Pandemic and Beyond. Acad Emerg Med. 2020 Nov;27(11):1183–6.

20. Chen C, Lee PI, Pain KJ, Delgado D, Cole CL, Campion TR. Replacing Paper Informed Consent with Electronic Informed Consent for Research in Academic Medical Centers: A Scoping Review. AMIA Jt Summits Transl Sci Proc. 2020 May 30;2020:80–8.

21. Richesson RL. Learning health systems, embedded research, and data standards— recommendations for healthcare system leaders. JAMIA Open. 2020 Oct 12;3(4):488– 91.

22. Gordon AL, Rick C, Juszczak E, Montgomery A, Howard R, Guthrie B, et al. The COVID-19 pandemic has highlighted the need to invest in care home research infrastructure. Age Ageing. 2022 Mar 1;51(3):afac052.

23. Bath PM, Ball J, Boyd M, Gage H, Glover M, Godfrey M, et al. Lessons from the PROTECT-CH COVID-19 platform trial in care homes. National Institute for Health and Care Research; 2025.

24. National Institute on Aging [Internet]. [cited 2025 Jun 16]. Recruitment: Electronic consent pilot. Available from: https://www.nia.nih.gov/research/milestones/translational-clinical-research/trial-innovation/milestone-12-e

25. Hammond M, Ashford P, High J, Clark LV, Howard G, Jones M, et al. Designing e-consent protocols for pragmatic clinical trials: case studies from a UKCRC clinical trials unit. Trials. 2024 Aug 19;25:550.

26. Kotting P, Beicher K, McKeith IG, Rossor MN. Supporting clinical research in the NHS in England: the National Institute for Health Research Dementias and Neurodegenerative Diseases Research Network. Alzheimers Res Ther. 2012 Jul 6;4(4):23.

27. Davies SL, Goodman C, Manthorpe J, Smith A, Carrick N, Iliffe S. Enabling research in care homes: an evaluation of a national network of research ready care homes. BMC Med Res Methodol. 2014 Apr 5;14:47.

28. Dementia TRC | NIHR [Internet]. 2024 [cited 2025 Jun 16]. Available from: https://www.nihr.ac.uk/about-us/what-we-do/infrastructure/translational-research-collaborations/dementia

29. Establishing a Clinical Trials Network [Internet]. Alzheimer’s Research UK. [cited 2025 Jun 16]. Available from: https://www.alzheimersresearchuk.org/about-us/our-influence/policy-work/reports/establishing-a-clinical-trials-network/

30. Join dementia research [Internet]. [cited 2025 Jun 16]. Available from: https://www.joindementiaresearch.nihr.ac.uk/

31. Inspire Fund – public engagement grant - Grant scheme [Internet]. Alzheimer’s Research UK. [cited 2025 Jun 16]. Available from: https://www.alzheimersresearchuk.org/grants/inspire-fund/

32. Personal and Public Involvement [Internet]. [cited 2025 Jun 16]. Available from: https://www.dementiasplatform.uk/about-us/dpuk-public-engagement

33. Angus DC, Alexander BM, Berry S, Buxton M, Lewis R, Paoloni M, et al. Adaptive platform trials: definition, design, conduct and reporting considerations. Nat Rev Drug Discov. 2019 Oct;18(10):797–807.

34. Response-adaptive randomization in clinical trials: from myths to practical considerations - PMC [Internet]. [cited 2024 May 29]. Available from: https://www.ncbi.nlm.nih.gov/pmc/articles/PMC7614644/

35. Pallmann P, Bedding AW, Choodari-Oskooei B, Dimairo M, Flight L, Hampson LV, et al. Adaptive designs in clinical trials: why use them, and how to run and report them. BMC Medicine. 2018 Feb 28;16(1):29.

36. Satlin A, Wang J, Logovinsky V, Berry S, Swanson C, Dhadda S, et al. Design of a Bayesian adaptive phase 2 proof-of-concept trial for BAN2401, a putative disease-modifying monoclonal antibody for the treatment of Alzheimer’s disease. Alzheimers Dement (N Y). 2016 Feb 4;2(1):1–12.

37. Wheatley A, Bamford C, Shaw C, Boyles M, Fox C, Allan L. Service organisation for people with dementia after an injurious fall: challenges and opportunities. Age and Ageing. 2019 May 1;48(3):454–8.

38. McKhann GM, Knopman DS, Chertkow H, Hyman BT, Jack CR, Kawas CH, et al. The diagnosis of dementia due to Alzheimer’s disease: Recommendations from the National Institute on Aging-Alzheimer’s Association workgroups on diagnostic guidelines for Alzheimer’s disease. Alzheimers Dement. 2011 May;7(3):263–9.

39. Care Home Research Network [Internet]. [cited 2020 Jul 8]. Available from: https://www.maudsleybrc.nihr.ac.uk/research/clinical-disorders/dementia/care-home-research-network/

40. Clinical Record Interactive Search (CRIS) [Internet]. [cited 2023 Mar 1]. Available from: https://www.maudsleybrc.nihr.ac.uk/facilities/clinical-record-interactive-search-cris/

