## supplementary materials for "Recruitment for dementia clinical trials in care homes: an evaluation of strategies employed in the Sativex® for the treatment of Agitation in Dementia (‘STAND’) Trial"

#### STAND Trial full eligibility criteria

**Inclusion Criteria**

- Aged 55-95 years
- Probable Alzheimer's disease per NINCDS-ADRDA criteria (38)
- Clinically significant agitation/aggression (CMAI ≥452 **and/or** NPI-NH Agitation ≥43) requiring treatment
- Nursing home residency with ≥2-week behavioral disturbance history
- Capacity-appropriate informed consent (participant, personal/professional legal representative)

**Exclusion Criteria**

Medication stability:

- Recent (≤2 weeks) dose changes: antipsychotics, antidepressants, benzodiazepines, hypnotics, lithium, or antiepileptics
- ChEIs/memantine dose changes within 6 weeks

Medical history:

- Current cannabis-based medications (UK-licensed)
- Concomitant strong CYP3A4 inducers/inhibitors
- Hypersensitivity to Sativex® components
- Severe cardiovascular disease (recent MI, uncontrolled hypertension, QTcF >450ms)
- Severe renal (eGFR <45 mL/min) or hepatic impairment (ALT/AST >3×ULN)
- Active delirium/pain-driven agitation
- Seizure history or fall risk (≥1 fall/6 months)

Psychiatric factors:

- Psychotic disorders/severe personality disorders
- Suicidality risk (C-SSRS >0)
- Substance/alcohol abuse history

Special populations:

- Females of childbearing potential
- Active COVID-19 infection (positive test ≤4 weeks/symptoms)

#### Comprehensive list of recruitment channels identified

1. **Care Home Research Network (CHRN)**: The CHRN connects over 200 care homes in London and southeast England to improve the quality of life for people with dementia. Convened by the NIHR Maudsley Biomedical Research Centre at King's College London, it fosters collaboration between care home workers and researchers, shaping scientific studies and sharing best practices (39).
2. **Dementia PPI Groups**: Patient and Public Involvement (PPI) groups comprise people living with dementia, their carers, and supporters who work as co-researchers and advisors in dementia research.
3. **CHRN Longitudinal Study**: Ongoing research within CHRN care homes that maintains cohorts of residents and staff. These cohorts can be approached for participation in additional studies.
4. **Care Home Intervention Team (CHIT)**: Specialist healthcare teams embedded in care homes who support residents' clinical needs. They help identify and facilitate recruitment of eligible participants.
5. **Community Mental Health Teams**: Multidisciplinary NHS teams providing mental health care to older adults, including those in care homes. They can identify and refer suitable participants for research.
6. **Clinical Records Interactive Service (CRIS)**: A secure NHS database that allows researchers to screen anonymised electronic health records for eligible participants. CRIS streamlines recruitment by enabling targeted identification (40).
7. **CQC Care Home Directory**: A public database listing all registered care homes in England, including contact details and service information. Researchers use it for direct outreach and targeted recruitment.
8. **GP Surgeries**: Primary care practices that maintain patient records and have ongoing relationships with people with dementia. GPs can identify and refer eligible patients to research studies.

9. **NIHR Clinical Research Networks**: National networks that support research delivery and participant recruitment across NHS and community settings. They provide research staff and established clinical links.
10. **Care Home Media and Podcasts**: Sector-specific publications and podcasts that reach care home professionals and decision-makers. These platforms are used to raise awareness and promote research opportunities.
11. **Social Media**: Online platforms used to share study information and engage with potential participants and carers. Social media enables broad outreach and community-driven recruitment, particularly Facebook which has a collection of privately led dementia care groups by members of the public.
12. **Mainstream Media**: General news outlets, such as newspapers and radio, that reach a wide public audience. These channels can increase awareness and interest in research participation.
13. **Join Dementia Research (JDR)**: A national registry matching volunteers interested in dementia research with suitable studies. It provides researchers with access to pre-screened, consented participants.
14. **Other Academic Departments**: University-based research teams with existing participant databases and collaborative networks. They can facilitate recruitment through cross-study referrals and academic partnerships.
15. **Memory Clinics**: Specialist NHS services diagnosing and managing dementia, maintaining clinically confirmed patient cohorts. Clinic staff can identify and refer eligible patients to research.
16. **Age-related charities**: Organisations such as the British Geriatrics Society, British Gerontological Society, Alzheimer's Research UK, Dementia UK, and the Care Provider Alliance represent healthcare professionals specialising in older adult care and provide networks for disseminating research opportunities. These societies offer access to clinicians working with target populations and can facilitate recruitment through professional networks, educational events, and by lending credibility and endorsement to research studies.
17. **Local Authorities**: Local government bodies overseeing social care and relationships with care providers. They can facilitate introductions and support research engagement in their jurisdictions.

#### Operationalising the priority matrix for clinical trial recruitment

In the context of clinical trials, stakeholders include patients, healthcare providers, collaborators, patient data sources, relevant media outlets, regulatory bodies, sponsors, and any other party that could feasibly have an impact or interest in the clinical trial you are conducting. Here's how I use Mendelow's Matrix to optimise recruitment in my clinical trials as a Clinical Trial Manager (CTM):

1. **Identify Stakeholders**: List all potential stakeholders involved in the clinical trial process. This includes patients, caregivers, healthcare professionals, regulatory authorities, funding bodies, media outlets/channels, patient registries/research cohorts, and any channel/group you can think of that may have an interest or influence on the work you are trying to do. It is crucial to think as far outside the box as you can - there are no wrong answers - there's no knowing which channel may be surprisingly effective in practice. Involve experts, clinicians, people with lived experience, advocacy groups etc.
2. **Assess Power and Interest**: Evaluate each stakeholder's power and interest concerning the clinical trial. For example, patients have high interest but varying levels of power, while patient registries have high power but may have lower day-to-day interest.
3. **Categorise Stakeholders**: Map each stakeholder into one of the four quadrants of Mendelow's Matrix. This process is dynamic throughout the trial, stakeholders will move between quadrants as their influence/interest rises and wanes. Ensure to regularly re-evaluate this step using ongoing recruitment analytics, PPI feedback, and mixed-methods research.
4. **Develop a strategic and tailored engagement strategy for each stakeholder group**:

**Key Players (Manage Closely)**: Engage patients and key healthcare providers with detailed information about the trial's benefits, risks, and procedures. Personalised communication and regular updates can build trust and encourage participation.

- *Role of the CTM*: Use ongoing trial recruitment analytics to monitor each key player channel to ensure they remain a 'key player'. Demote and reallocate resources as required if key player recruitment channels 'dry up'. Delegate responsibility to proactive trial team members to ensure these key players are always catered for. 'Titrate' engagement as required - start with email, then phone call, and meet in-person if needed. Once potential participants are identified & engaged, develop & implement a standardised consenting algorithm. This algorithm should detail a every potential pathway to a participant consenting, or not consenting. The ultimate goal is to achieve an outcome either way - close the loop! It should detail available methods of consent, who can provide consent, how long to wait for response, when/whether to move on to other proxies' etc. Ensure to train every member of the trial team on this process.

**Keep Satisfied**: Provide high-level updates to regulatory bodies and sponsors. Ensure compliance and address any concerns promptly to maintain their support.

- *Role of the CTM*: This is even more important than managing key players (which can be delegated to trial team members). The primary objective of the CTM for this stakeholder group is to increase their interest with whatever methods are most appropriate and tailored to the specific group. The idea is to move as many 'keep satisfied' stakeholders into a 'key player' category as possible so that their influence can be maximised toward the success of the trial (and trial recruitment!).

**Keep Informed**: Share regular updates with patient advocacy groups and community organisations. Their support can help spread awareness and encourage participation.

- *Role of the CTM*: When possible, collaborate with these stakeholders to share their high interest to increase the interest of 'Keep Satisfied' stakeholders, moving them to key players. In doing so, many will also start to increase their own influence on the study, and move toward 'Keep Satisfied'.

**Monitor**: Keep an eye on stakeholders with low power and interest, such as peripheral healthcare providers. At the minimum, ensure they have basic information and remain neutral or supportive.

*Role of the CTM:* increase the level of interest of these stakeholders, so that they move into the 'keep informed' category. Prioritise other stakeholder quadrants first though.

### STAND Trial consenting protocol flow diagram.

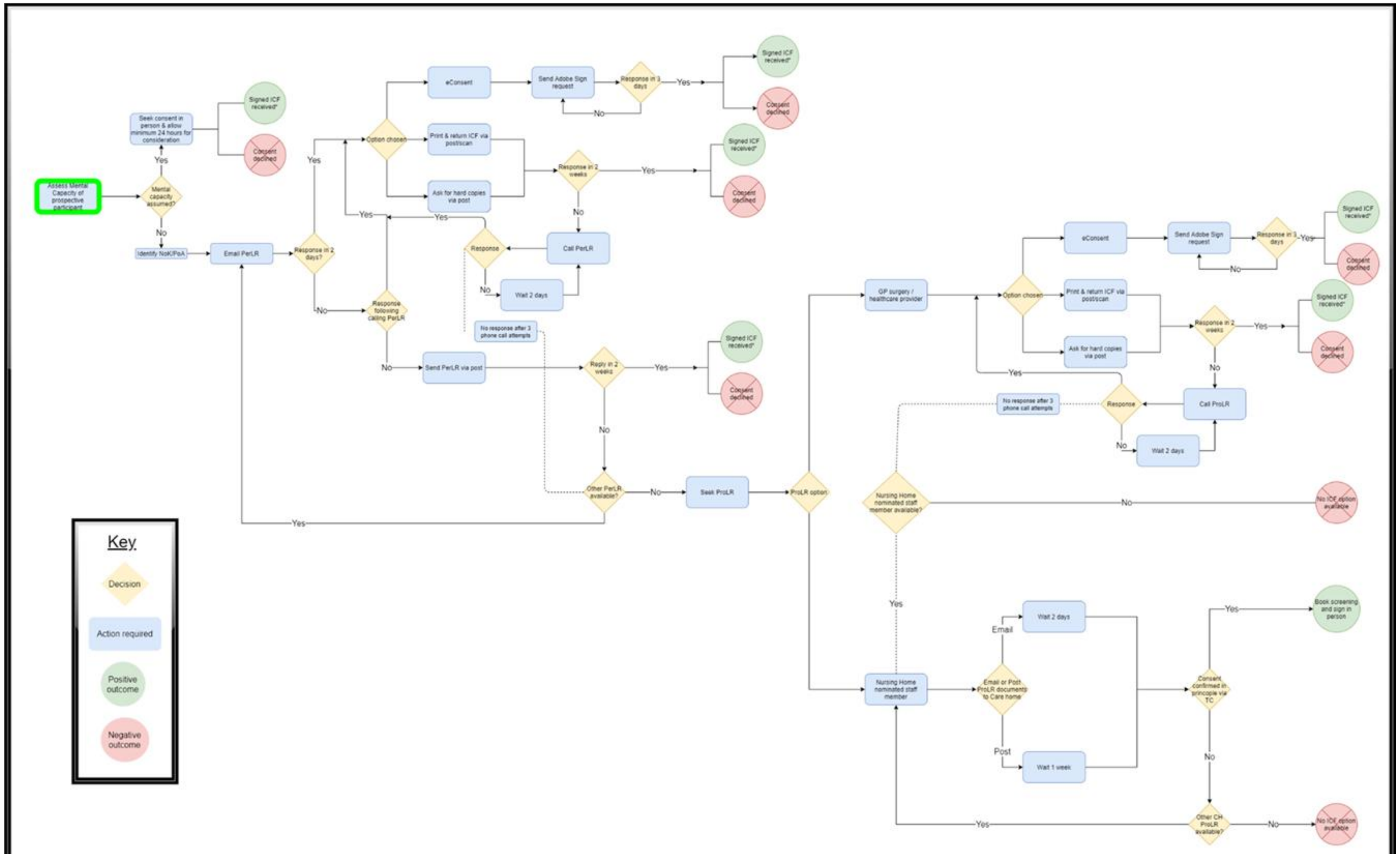


**Abbreviations**: NoK/PoA - Next of Kin/Power of Attorney; PerLR - Personal Legal representative; ProLR - Professional Legal Representative; ICF - Informed Consent Form; CH - Care Home;
*Receipt of Informed consent form **does not** mean participant has consented to participate; instead it simply reflects that a decision has been made either way.
